# Unraveling Metabolic Signatures in Breast Cancer: Machine Learning for Improved Therapeutic Targeting

**DOI:** 10.1101/2025.03.18.25324194

**Authors:** Fatemeh Mehdikhani, Parnian Habibi, Sajad Alavimanesh

## Abstract

**Background:** Breast cancer is one of the leading causes of cancer-related mortality among women worldwide. Despite advancements in treatment, therapeutic resistance remains a major challenge, necessitating novel approaches for more effective interventions. One of the hallmarks of cancer, particularly in breast tumors, is metabolic reprogramming, where altered metabolic pathways create distinct profiles compared to normal cells. Identifying these metabolic alterations can provide critical insights for developing targeted therapies aimed at disrupting tumor metabolism and improving patient outcomes.

**Objectives:** This study applies six machine learning algorithms to predict metabolic profiles in breast cancer patients compared to healthy individuals, providing a promising approach for identifying metabolic targets in precision therapy.

**Method:** Plasma samples from 102 breast cancer patients and 99 healthy individuals were analyzed using targeted liquid chromatography-tandem mass spectrometry (LC-MS/MS) to assess metabolic profiles. Six machine learning algorithms were applied to evaluate classification performance, and feature importance was determined using the Mean Squared Error (MSE) value.

**Result:** Our findings revealed a significant decrease in alanine, histidine, tryptophan, tyrosine, methionine, and proline levels in breast cancer patients. Among the machine learning models, Random Forest (RF) achieved the highest classification performance (accuracy: 0.90, specificity: 0.85, sensitivity: 0.95), followed by K-Nearest Neighbors (KNN) with similar sensitivity but lower specificity. Logistic Regression (LR) balanced specificity (0.90) and sensitivity (0.86) with an accuracy of 0.88. Naïve Bayes (NB) and Support Vector Machine (SVM) showed moderate accuracy (0.83), while Decision Tree (DT) had the lowest sensitivity (0.76) but the highest PPV (0.89). Feature importance analysis identified glutamic acid, ketocholesterol, cystine, ornithine, succinate, acetylcarnitine, asparagine, tryptophan, and palmitic acid as key metabolic markers.

**Conclusion:** This study draws attention to key predictive metabolic bottlenecks identified through machine learning models, which could aid in targeted therapy and personalized treatment based on patients’ metabolic profiles.

## 1. Introduction

Breast cancer is one of the most prevalent malignancies among the female population worldwide, accounting for a significant portion of cancer-related morbidity and mortality (1). Despite advancements in treatment strategies, overcoming therapeutic resistance and improving patient outcomes remain critical challenges (2). In recent years, cancer metabolism has gained considerable attention, particularly due to the Warburg effect, a phenomenon in which cancer cells preferentially utilize glycolysis for energy production, even in the presence of oxygen(3). This metabolic shift supports rapid cell proliferation and survival under various stress conditions, a process known as metabolic reprogramming (4). Given its fundamental role in tumor progression, metabolism has emerged as a defining characteristic of cancer cells (5).

Several metabolic pathways are altered in breast cancer. Glycolysis is highly active, accompanied by elevated expression of glucose transporters and glycolytic enzymes (6, 7). Beyond glucose metabolism, specific amino acids, such as serine and glutamine, are essential for breast cancer growth (8, 9). Additionally, increased activation of the pentose phosphate pathway (PPP) has been reported in breast tumors, contributing to nucleotide biosynthesis and redox homeostasis (10). Moreover, lipid metabolism is dysregulated, with breast cancer cells utilizing fatty acids and lipid synthesis, including cholesterol uptake, to sustain membrane biosynthesis and energy production (11). These metabolic adaptations enable cancer cells to thrive in hostile microenvironments and contribute to tumor progression (12).

Predictive medicine is an emerging field that leverages advanced technologies such as genomics, bioinformatics, and artificial intelligence to assess disease risk and personalize healthcare (13-15). By analyzing genetic variations, lifestyle factors, and clinical data, predictive models can identify individuals susceptible to diseases like diabetes, cancer, and cardiovascular disorders, allowing for early diagnosis and targeted prevention strategies (16-18). Machine learning algorithms enhance predictive accuracy by detecting complex patterns in large datasets, enabling personalized treatment plans and improving patient outcomes (19-21). Additionally, pharmacogenomics, a key component of predictive medicine, tailors drug therapies based on an individual’s genetic makeup, minimizing adverse reactions and optimizing efficacy (22-25). As predictive medicine continues to evolve, it is reshaping healthcare by shifting the focus from reactive treatment to proactive prevention and precision care (26-28).

Given the distinct metabolic alterations between breast tumors and normal tissues, metabolic profiling holds great potential for improving breast cancer treatment strategies. Identifying these metabolic vulnerabilities may provide novel therapeutic targets, serving as metabolic bottlenecks that can be exploited to hinder tumor progression and enhance treatment efficacy. This study aims to investigate metabolic signatures in breast cancer patients and assess their clinical relevance using machine learning approaches. By analyzing metabolic profiles, we hypothesize that distinct metabolic patterns can serve as predictive metabolic bottlenecks for breast cancer, ultimately contributing to the development of more effective and personalized treatment strategies.

## 2. Methods

### 2.1. Study Design and Participants

This study utilized a dataset comprising demographic and metabolic data from 201 individuals, including 102 breast cancer patients and 99 healthy controls. The study adhered to the principles of the Declaration of Helsinki and was approved by the Inonu University Health Sciences Non-Interventional Clinical Research Ethics Committee (protocol code = 2024/5750) for data analysis. Prior to sample collection, informed consent was obtained from all participants in accordance with ethical guidelines. Blood samples were collected following an overnight fasting period to minimize metabolic variability. Plasma metabolic profiles were then analyzed using targeted liquid chromatography-tandem mass spectrometry (LC-MS/MS) to ensure precise quantification of metabolites. After data acquisition, the samples were categorized based on demographic characteristics and key metabolic markers for further analysis (29, 30).

### 2.2. Sample Preparation and LC-MS/MS Analysis

Plasma samples were thawed overnight at 4□°C before metabolite extraction. For each sample, 50□μL of plasma was transferred to a 2□mL vial, followed by protein precipitation using 300□μL of methanol. The mixture was vortexed, incubated at -20□°C, and subjected to sonication and centrifugation. The supernatant was collected, dried using a vacuum concentrator, and reconstituted in a solvent containing stable isotope-labeled internal standards for system performance monitoring. A pooled quality control (QC) sample, composed of plasma from both breast cancer patients and healthy controls, was processed identically and analyzed at regular intervals. LC-MS/MS experiments were conducted using a Waters Acquity I-Class UPLC TQS-micro MS system. Chromatographic separation was performed on a Waters Xbridge BEH Amide column at 40□°C with a flow rate of 0.3□mL/min. Samples were analyzed in both positive and negative ionization modes, using distinct mobile phase compositions for each mode. A gradient elution program was applied to optimize metabolite separation, and metabolite identities were confirmed by spiking standard compounds into plasma samples. Data acquisition and integration of extracted multiple reaction monitoring (MRM) peaks were performed using TargetLynx software (29).

### 2.3. Study Measures

#### Metabolic Markers

Plasma metabolites were analyzed using LC-MS/MS, targeting key metabolic pathways associated with breast cancer progression.

#### Other Clinical Variables

Age was included as a covariate in statistical models.

### 2.4. Statistical Approach

#### 2.4.1. Machine Learning Models

A machine learning-based classification approach was applied to metabolic data to categorize individuals into breast cancer patients and healthy controls. Six different machine learning models were evaluated:

##### 2.4.1.1. K-Nearest Neighbors (KNN)

K-Nearest Neighbors (KNN) is a machine learning algorithm that classifies data points based on their proximity to neighboring samples. In this distance-based model, a new sample is categorized according to the most frequent class among its k nearest neighbors (31).

##### 2.4.1.2. Support Vector Machine (SVM)

Support Vector Machine (SVM) is a machine learning algorithm used for data classification. It constructs a decision boundary, known as a hyperplane, to separate different classes and assigns data points to their respective categories. By maximizing the margin between classes, SVM minimizes overfitting and enhances generalization, making it useful for handling complex data structures (32).

##### 2.4.1.3. Naïve Bayes (NB)

Naïve Bayes is a statistical classification algorithm that predicts class membership probabilities. It estimates the probability of a sample being assigned to a particular class, assuming that each attribute contributes to the classification independently of the others. Naïve Bayes is based on Bayes’ Theorem (33).

##### 2.4.1.4. Decision Tree (DT)

A decision tree is a supervised learning method that classifies data using a tree-like structure consisting of a root node, branch nodes, and leaf nodes. The dataset is recursively divided into smaller subsets, and the decision tree is built incrementally to improve classification accuracy.

##### 2.4.1.5. Random Forest (RF)

The Random Forest algorithm constructs multiple decision trees and classifies data based on a majority vote. Each decision tree is trained on a different subset of the data, reducing the risk of overfitting. This method enhances model stability and improves accuracy, making Random Forest a reliable and robust classification algorithm.

##### 2.4.1.6. Logistic Regression (LR)

Logistic regression (LR) is a supervised classification algorithm that predicts the probability of an outcome based on continuous or categorical independent variables. It is useful for determining the presence or absence of a characteristic and assumes independent sampling and inclusion of all relevant predictors. As a linear model, LR establishes a weighted relationship between independent variables and the target class.

#### 2.4.2. Model Training and Evaluation

The classification models were trained to distinguish between breast cancer patients and healthy individuals. The dataset was randomly split into training (80%) and testing (20%) subsets. Model performance was assessed using the following metrics: Accuracy, Sensitivity (True Positive Rate), Specificity (True Negative Rate), Positive Predictive Value (PPV), and Negative Predictive Value (NPV). To ensure robustness and prevent overfitting, a 5-fold cross-validation strategy was applied.

#### 2.4.3. Statistical Analysis

All statistical analyses were conducted using R software (version 4.4). Continuous variables with a normal distribution were analyzed using an independent t-test for comparison, with a significance threshold set at p < 0.001.

## Results

### 3.1. Sample Characteristics

The demographic and certain significant metabolic characteristics of the participants are summarized in Table 1. A total of 201 plasma samples were analyzed, including 102 samples from breast cancer patients and 99 from healthy controls. The average age of breast cancer patients was 55 years, while that of healthy individuals was 52 years. Additionally, the mean plasma metabolite levels were measured across participants. For instance, Alanine levels were 377036 in patients versus 519367 in healthy individuals, and Histidine levels were 1080024 in patients compared to 1210992 in healthy individuals, Tryptophan levels were 978986 in patients compared to 1196444 in healthy individuals. A comprehensive analysis of all metabolic data can be found in Supplementary Table 1.

**Table 1.** Sample Characteristics.

|  | Healthy Samples<br>Mean (SD) | Breast cancer patients<br>Mean (SD) | p |
| --- | --- | --- | --- |
| Age | 52 (12) | 55 (10) | 0.056 |
| Alanine | 519367 (141095) | 377036 (121443) | <0.001 |
| Histidine | 1210992 (239513) | 1080024 (241916) | <0.001 |
| Tryptophan | 1196444 (393159) | 978986 (291736) | <0.001 |
| Acetylcarnitine | 293355 (167744) | 492791 (225758) | <0.001 |
| Acetylglucosamine | 3440 (1187) | 2668 (1054) | <0.001 |
| Adenosine | 1316 (1224) | 2646 (3235) | <0.001 |
| Tyrosine | 113269 (43105) | 76600 (24208) | <0.001 |
| Anthranilic.acid | 907 (652) | 584 (546) | <0.001 |
| Caffeine | 390620 (423610) | 104374 (182222) | <0.001 |
| Carnitine | 1466179 (347252) | 1266450 (344290) | <0.001 |
| Choline | 832622 (250867) | 684913 (258019) | <0.001 |
| Creatinine | 8469468 (1899658) | 6979915 (1742566) | <0.001 |
| Cystine | 60124 (48938) | 138303 (46830) | <0.001 |
| Glutamic Acid | 1253120 (695248) | 412072 (216153) | <0.001 |
| Methionine | 151370 (67815) | 193134 (57982) | <0.001 |
| Homoserine | 514689 (147898) | 409621 (122108.97) | <0.001 |
| Hypoxanthine | 193253 (261932) | 402543 (428699) | <0.001 |
| Isoleucine | 6439637 (2162432) | 4789288 (1186628) | <0.001 |
| Kynurenic.acid | 3356 (1581) | 2307 (1282) | <0.001 |
| Kynurenine | 11791 (4565) | 9463 (3686) | <0.001 |
| L.Alloisoleucine | 7108170 (2376444) | 5287973 (1312879) | <0.001 |
| Leucine | 6414989 (2157010) | 4757621 (1186610) | <0.001 |
| Lysine | 1222998 (340572) | 975897 (311635) | <0.001 |
| Proline | 621213 (202214) | 438417 (170220) | <0.001 |
| Norleucine | 5959529 (1997544) | 4423950 (1098147) | <0.001 |
| Ornithine | 595850 (220847) | 341889 (133200) | <0.001 |
| Phenylalanine | 3274991 (993388) | 2463481 (551597) | <0.001 |

### 3.2. Classification Performance

The performance of six machine learning models: K-Nearest Neighbors (KNN), Support Vector Machine (SVM), Naïve Bayes (NB), Decision Tree (DT), Random Forest (RF), and Logistic Regression (LR) was evaluated using sensitivity, specificity, accuracy, positive predictive value (PPV), and negative predictive value (NPV). The detailed results are summarized in Figure 1. Among the classifiers, RF achieved the highest accuracy (0.90) and specificity (0.85), with sensitivity equal to KNN (0.95). KNN also demonstrated high sensitivity (0.95) and a strong NPV (0.94), though its specificity was lower (0.75). LR showed a balance between specificity (0.90) and sensitivity (0.86) with an overall accuracy of 0.88. NB and SVM achieved similar accuracy scores (0.83), with NB showing slightly higher sensitivity (0.90) but lower specificity (0.75). The DT model had the lowest sensitivity (0.76) but the highest PPV (0.89), indicating strong performance in positive class prediction. Overall, RF demonstrated the highest classification performance, followed by LR and KNN, based on accuracy and sensitivity.

**Figure 1.**
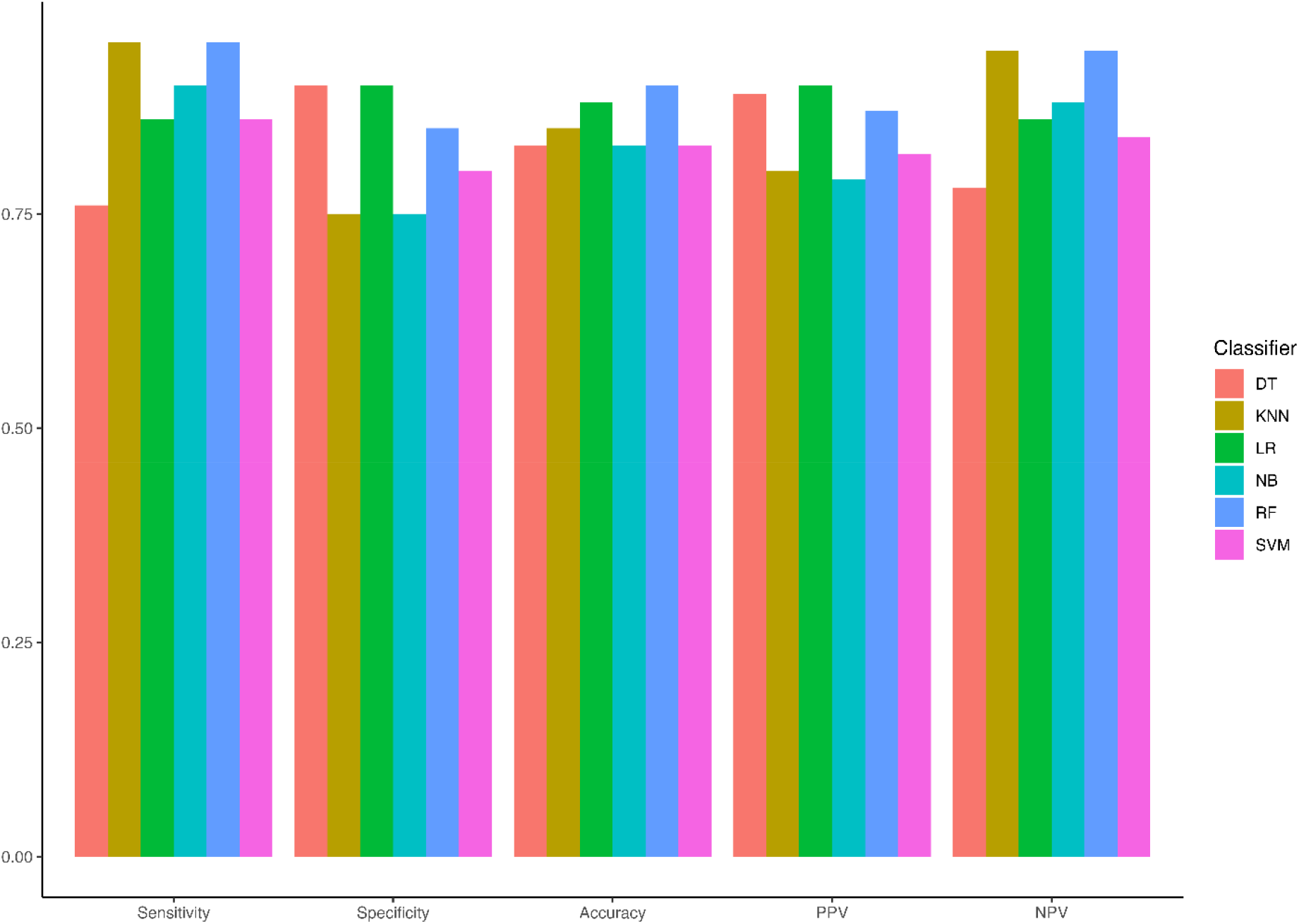
Performance Metrics of Machine Learning Models on the Test Dataset

### 3.3. Feature Importance Analysis

To determine the most influential features in classification performance, the Percentage Increase in Mean Squared Error (%IncMSE) was used as the importance metric. Figure 2 presents the ranking of key features based on this measure, identifying the top 10%. Among them, glutamic acid (16.31) exhibited the highest value, emphasizing its strong contribution to model accuracy. Other significant features within the top 10% included ketocholesterol (12.45), cysteine (10.60), age (10.49), ornithine (6.65), succinate (5.96), acetylcarnitine (5.96), asparagine (5.38), tryptophan (5.25), and palmitic acid (5.24), all of which played a crucial role in model performance.

**Figure 2.**
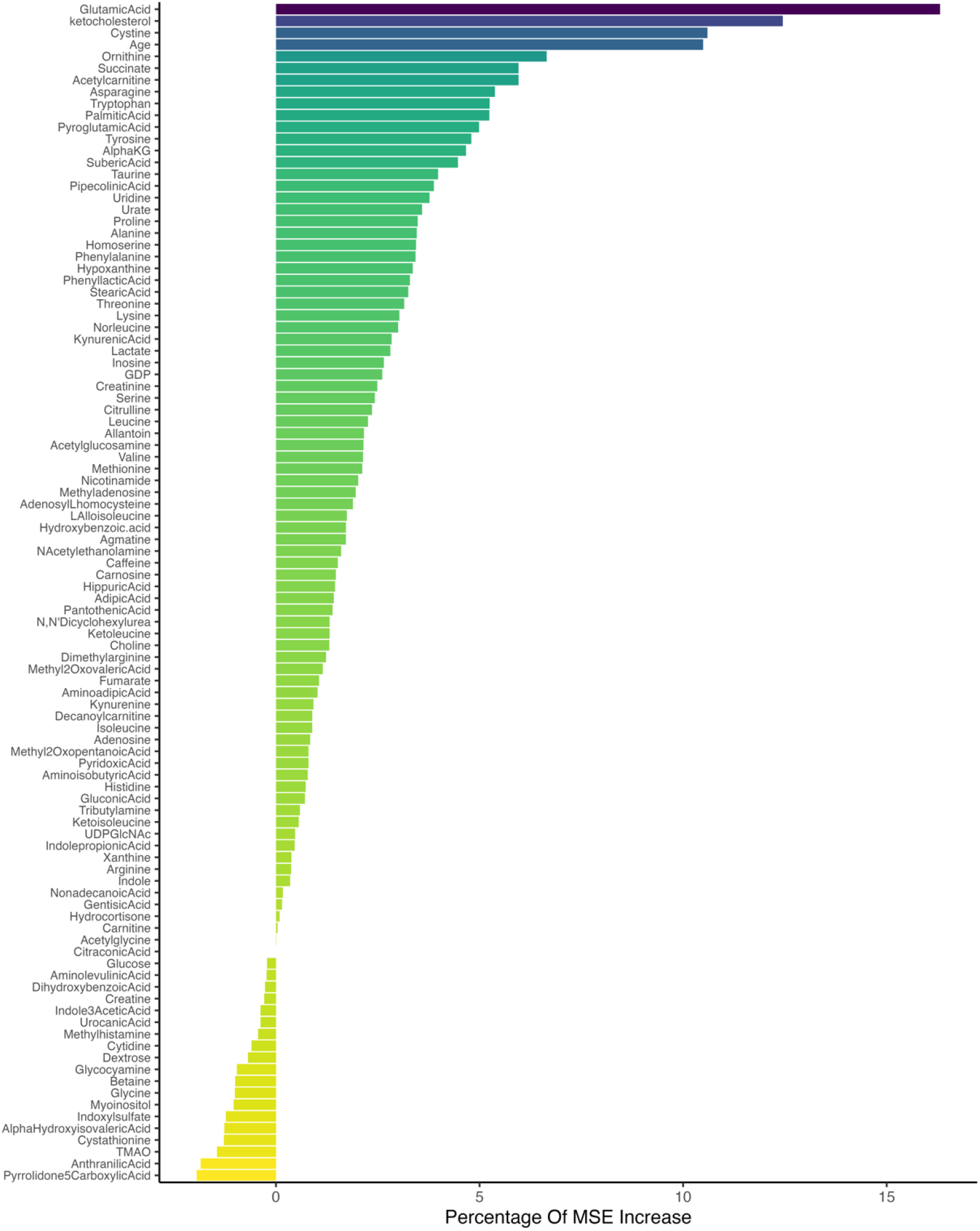
Ranking of the Most Important Features Based on %IncMSE

## 4. Discussion

In this study, we explored metabolic profiling as a potential tool for distinguishing breast cancer patients from healthy individuals. Given that metabolism is a hallmark of cancer, our aim was to identify metabolic signatures that could serve as biomarkers for breast cancer detection and potential therapeutic targets. We identified distinct metabolic alterations in the plasma samples of breast cancer patients compared to healthy individuals. Notably, the concentration of several amino acids, including alanine (Ala), histidine (His), tryptophan (Trp), tyrosine (Tyr), methionine (Met), and proline (Pro), was significantly reduced in breast cancer patients. Using machine learning models, we successfully classified individuals based on their metabolic profiles. Among the models tested, RF achieved the highest accuracy (0.90) and specificity (0.85), while KNN exhibited the highest sensitivity (0.95). The high sensitivity of RF and KNN suggests their potential for detecting cancer cases with minimal false negatives, which is crucial for early diagnosis. However, KNN had a higher false-positive rate (specificity: 0.75), which may lead to unnecessary follow-ups. LR showed a well-balanced performance (accuracy: 0.88, specificity: 0.90, sensitivity: 0.86), making it a suitable choice when minimizing both false positives and false negatives. In contrast, NB and SVM performed moderately (accuracy: 0.83), with NB favoring sensitivity (0.90) at the cost of specificity (0.75), making it more suitable for screening applications where missing positive cases is a greater concern than false positives. Feature importance analysis highlighted glutamic acid, 7-ketocholesterol (7-KC), cystine, age, ornithine, succinate, acetylcarnitine, asparagine, tryptophan, and palmitic acid as the most significant contributors to model accuracy.

Our findings align with previous research demonstrating altered amino acid metabolism in breast cancer. We observed a significant decrease in plasma levels of Ala, His, Trp, Tyr, Met, and Pro in breast cancer patients, consistent with a study by Shen et al., which reported reduced levels of these amino acids in triple-negative breast cancer (TNBC) patients. Additionally, in ER+/PR+ breast cancer patients, levels of Ala and His were significantly lower than in healthy controls (34). Another study analyzing plasma from patients with luminal A, TNBC, and HER2-positive breast cancer also observed a significant decrease in Trp levels compared to healthy individuals (35). These findings suggest that amino acid metabolism is systematically altered in breast cancer, potentially due to increased tumor cell consumption and metabolic reprogramming.

Feature importance analysis identified glutamic acid as one of the most significant metabolic markers in breast cancer. Glutamic acid is a central metabolite in cancer metabolism, particularly in the glutaminolysis pathway, which fuels tumor growth by supplying carbon and nitrogen sources for biosynthesis and redox balance maintenance (36, 37). Studies indicate that glutaminase (GLS), the enzyme converting glutamine to glutamic acid, is upregulated in aggressive breast cancers such as TNBC, promoting proliferation and survival Additionally, increased glutamic acid levels have been linked to therapy resistance in endocrine-resistant breast cancer (38). Given these roles, targeting glutaminase or glutamic acid metabolism could represent a promising therapeutic strategy, particularly in cancers that exhibit glutamine addiction.

Our study also identified 7-KC as a key metabolic feature in breast cancer. 7-KC is an oxidized cholesterol derivative implicated in cancer progression and drug resistance (39). Research suggests that 7-KC reduces doxorubicin cytotoxicity in ER+ MCF-7 cells by upregulating P-glycoprotein via an ERα- and mTOR-dependent pathway, leading to reduced intracellular drug accumulation and decreased efficacy (39). Furthermore, 7-KC has been shown to modulate tamoxifen response, slightly reducing its effect in ER+ cells while enhancing it in ER-negative BT-20 cells. It also promotes cancer cell migration and invasion, suggesting a role in breast cancer metastasis (40). However, recent studies indicate that 7-KC-loaded phosphatidylserine liposomes exhibit anticancer potential by inducing apoptosis and autophagy in melanoma and breast adenocarcinoma models, highlighting its dual role in cancer biology (41). Overall, 7-KC influences both drug resistance and tumor progression, underscoring the need for further research into its therapeutic potential. Cystine, the oxidized dimer of cysteine, plays a crucial role in breast cancer progression, particularly in TNBC (42). Our study identified cystine as a significant metabolic marker, aligning with studies showing that TNBC cells, especially mesenchymal subtypes, exhibit a strong dependency on cystine for survival. TNBC cells are highly sensitive to cystine deprivation, which induces programmed necrosis through TNFα and the MEKK4-p38-Noxa pathways. Interestingly, inhibiting these pathways reduces cell death, suggesting potential therapeutic strategies targeting cystine metabolism in aggressive breast cancer subtypes (42).

## Limitations

Despite the promising findings of our study, several limitations should be acknowledged. First, our sample size may not fully capture the heterogeneity of breast cancer, and larger, multi-center studies are needed to validate the metabolic signatures identified. Second, while we used machine learning models for classification, further optimization and external validation are required to confirm their clinical applicability. Incorporating additional datasets from independent cohorts could improve model generalizability. Third, our study relied on plasma metabolomics, which provides valuable insights but does not directly reflect tumor-specific metabolic changes. Integrating tumor tissue metabolomics or single-cell analysis could offer a more comprehensive understanding of metabolic alterations. Finally, although we identified key metabolic features linked to breast cancer, functional studies are needed to elucidate their precise mechanistic roles in tumor progression and therapy resistance.

## Conclusion and future directions

This study highlights the potential of metabolic profiling and machine learning in breast cancer detection and classification. Our findings reveal significant alterations in amino acid metabolism, with decreased levels of alanine, histidine, tryptophan, tyrosine, methionine, and proline in breast cancer patients compared to healthy individuals. Feature importance analysis identified glutamic acid, 7-KC, and cystine as key metabolic markers, providing new insights into cancer metabolism and therapeutic targeting. Among the machine learning models tested, RF demonstrated the highest classification performance, followed by LR and KNN. These findings highlight the potential of metabolic markers and machine learning approaches in advancing non-invasive breast cancer diagnostics. Future studies should focus on validating these findings in larger cohorts, integrating multi-omics approaches, and exploring the mechanistic role of identified metabolites in breast cancer progression and treatment resistance.

## Data Availability

All data produced in the present study are available upon reasonable request to the authors

## Abbreviation

mTOR: mechanistic Target of Rapamycin
TNFα: Tumor Necrosis Factor Alpha
MEKK4: Mitogen-Activated Protein Kinase Kinase Kinase 4
TNBC: Triple-Negative Breast Cancer
KNN: K-Nearest Neighbors
NB: Naïve Bayes
SVM: Support Vector Machine
DT: Decision Tree
RF: Random Forest
LR: Logistic Regression

## Ethics statement

This study was approved by the Inonu University Health Sciences Non-Interventional Clinical Research Ethics Committee and conducted in compliance with institutional guidelines, local regulations, and the ethical principles outlined in the Declaration of Helsinki. Prior to participation, all individuals provided written informed consent.

## Author Contributions

F.M., Conceptualization, Formal Analysis, Methodology, Software, Visualization, Writing–original draft, Writing–review and editing, P.H., Conceptualization, Formal Analysis, Methodology, Software,Visualization, S.A., Writing–original draft, Writing– review and editingwriting—review and editing. All authors have read and agreed to the published version of the manuscript.

